# Evaluation of the causal associations between brain imaging-derived phenotypes and type 2 diabetes: a bidirectional Mendelian randomization study

**DOI:** 10.1101/2022.12.16.22283558

**Authors:** Shan-Shan Dong, Ke Yu, Shi-Hao Tang, Jing Guo, Yan Guo, Tie-Lin Yang

**Author notes:** Corresponding authors: Tie-Lin Yang, Ph.D., Yan Guo, Ph.D.

## Abstract

**OBJECTIVE:** To investigate whether the association between brain imaging-derived phenotypes (IDPs) and Type 2 diabetes (T2D) related traits is causal.

**DESIGN:** Two sample, bidirectional Mendelian randomization study.

**SETTING:** Genome wide association study (GWAS) summary data taken from various cohorts comprised of the general population (mainly composed of Europeans).

**PARTICIPANTS:** Summary data were used from previous GWAS. For IDPs, the data included up to 33,224 European individuals from the UK Biobank. For T2D-related traits, the number of participants ranged from 63,396 to 455,313.

**MAIN OUTCOME MEASURES:** A total of 587 reliable IDPs and five T2D-related traits (T2D, fasting glucose, 2h-glucose post-challenge, glycated hemoglobin, and fasting insulin).

**RESULTS:** We identified 3 IDPs with potential causal effects on T2D or fasting insulin. For example, we observed that the area of the right rostral middle frontal cortex was negatively associated with the T2D risk (OR = 0.74, 95% CI 0.65 to 0.85, *P* = 1.31 × 10^−5^). In addition, we identified potential causal effects of T2D-related traits on 6 IDPs. For example, T2D was negatively associated with the volumes of the right superior frontal gyrus (β = -0.05, 95% CI -0.08 to -0.03, *P* = 2.17 × 10^−5^) and the right paracentral lobule (β = -0.05, 95% CI -0.07 to -0.02, *P* = 1.74 × 10^−4^).

**CONCLUSIONS:** Our results revealed strong genetic evidence for the bidirectional causal associations between brain neuroimaging phenotypes and T2D-related traits. This will contribute to better prediction and intervention for the risk of T2D.

## Introduction

Type 2 diabetes (T2D) is a metabolic disorder that is characterized by relative insulin deficiency, insulin resistance, and hyperglycemia. T2D has become a global pandemic in recent decades. In 2017, approximately 462 million individuals were affected by T2D [1]. Concerningly, the T2D incidence continues to increase, it is estimated that over 590 million T2D patients will be diagnosed by 2035 [2]. Elucidating the potential causal factors and consequences for T2D might be beneficial for informing prevention strategies.

Historically, pancreatic islet has been viewed as principal regulators of glycemia. However, accumulating evidence suggests that the brain cooperates with the islet to control glucose homeostasis [3]. The brain can regulate islet function directly via the autonomic nervous system and indirectly via neuroendocrine mechanisms [4]. Interventions targeting the brain could regulate insulin sensitivity and glucose metabolism [5]. On the other hand, the relationship between T2D and the brain maybe bidirectional. T2D was reported as a risk factor for cognitive impairment and cerebrovascular disease [6-8]. The pathogenesis is still unclear, but chronic hyperglycemia and possibly direct effects of insulin on the brain have been implicated [9, 10]. Therefore, it is imperative for research to understand the pathways that link brain function and T2D.

Brain imaging, which measures the living brain in a non-invasive way using magnetic resonance imaging [11], can help to clarify the pathways linking the brain and T2D. Many observational studies have been performed to explore the relationships between brain imaging-derived phenotypes (IDPs) and T2D. Several studies reported reduced total brain volume in T2D patients [12, 13]. T2D were associated with greater white matter hyperintensity (WMH) in some [14, 15] but not all studies [16, 17]. The findings for the associations between T2D and brain sub-regions are also inconsistent. For example, some studies [16] reported that T2D was associated with lower hippocampal volumes while others [12] found no association. These studies highlight the ongoing debate about the associations between brain structure and T2D. Moreover, results from observational studies are unable to fully account for confounding factors. Therefore, whether the relationship between T2D and IDPs is causal is uncertain.

Mendelian randomization (MR), which uses genetic markers associated with the exposure as instruments, has been widely used to assess the causal relationship between exposure and outcome [18]. Since genetic variants are randomly allocated at meiosis and fertilization, they are independent of confounding factors or reverse causation that might bias observational studies. Two-sample MR analysis [19], which extracted instrument variables from summary statistics of large-scale non-overlapping genome-wide association studies (GWAS) has been widely used to explore the causal relationships between interested exposure and outcome. In this study, we performed bidirectional two-sample MR analyses to estimate the causal associations between 587 IDPs and five T2D-related traits (T2D, fasting glucose (FG), 2h-glucose post-challenge (2hGlu), glycated hemoglobin (HbA1c), and fasting insulin (FI)). Our results could help to clarify the causal associations between IDPs and T2D, which might offer further insight into the pathophysiology of T2D and guide research efforts toward disease prevention.

## Methods

This study is reported in accordance with the Strengthening the Reporting of Observational Studies in Epidemiology (STROBE) guideline [20]. We used deidentified publicly available data, so the ethical approval from an institutional review board was not was required.

### Data sources

#### Brain IDPs

GWAS summary data for the brain IDPs were generated by Smith *et al*.[21], including up to 33,224 European individuals from the UK Biobank. We downloaded the data from the Oxford Brain Imaging Genetics web browser (https://open.win.ox.ac.uk/ukbiobank/big40/) [21]. Referring to our previous study [22], we selected 587 reliable and meaningful brain structural IDPs from the original releases, and these IDPs were further classified into 13 regional categories and 9 measures (Figure 1). When IDPs were used as exposures, the reported effect of MR analyses is based on per standard deviation (SD) changes of the IDPs. For each IDP, we calculated the SD using the individual-level brain imaging data from UK Biobank under the data application ID 46387. The calculation was mainly based on the data collected in 2019. If data in 2019 were not available, the data measured in 2014 were used.

**Figure 1.**
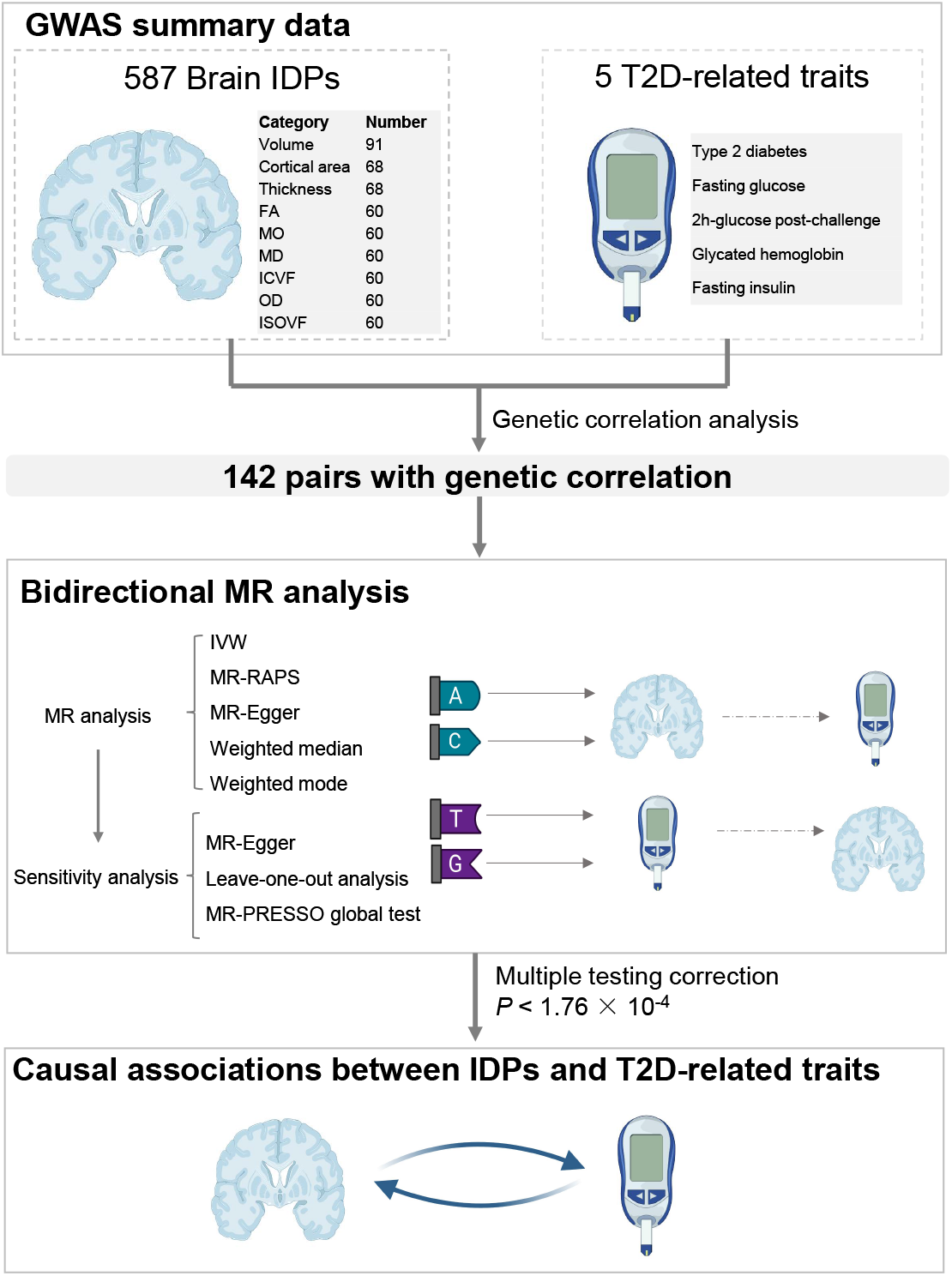
Outline of the current study. Abbreviations: IDP: Imaging-derived phenotype; FA: fractional anisotropy; MD: mean diffusivity; MO: diffusion tensor mode; ICVF: intracellular volume fraction; ISOVF: isotropic or free water volume fraction; OD: orientation dispersion index.

#### T2D-related traits

We used 5 T2D-related traits in our analyses, including T2D, fasting glucose (FG), 2h-glucose post-challenge (2hGlu), glycated hemoglobin (HbA1c), and fasting insulin (FI). The GWAS summary data for T2D was generated by Mahajan *et al*., [23], we downloaded the dataset without UK Biobank subjects from the DIAGRAM (DIAbetes Genetics Replication and Meta-analysis) consortium web site (https://diagram-consortium.org/downloads.html). The summary data for the rest four traits were generated by Chen *et al*. [24],we obtained the datasets including European only subjects from the MAGIC (Meta-Analyses of Glucose and Insulin-related traits Consortium) web site (https://magicinvestigators.org/downloads/). Detail information of these datasets is shown in Supplementary Table S1.

### Genetic correlation analyses

If two traits with non-zero heritability are causally related, there should be genetic correlations between them [25]. Therefore, prior to MR analysis, we performed genetic correlation analysis between IDPs and T2D-related traits using LD score regression [26] with default parameters. IDPs potentially associated with T2D-related traits were subsequently subjected to further MR analysis. For genetic correlation analysis, we used *P* < 0.05 as the threshold to preserve all datasets with suggestive evidence.

### MR analyses

#### Instrument variants (IV) selection

For each exposure, we used the clumping algorithm in PLINK (version 1.9) [27] to select independent SNPs (r^2^ threshold = 0.001, window size = 1 Mb and *P* < 5 × 10 ^−8^) as IVs. The 1000G European data (phase 3) were used as the reference for LD estimation. SNPs with minor allele frequency < 0.01 or SNPs within long LD regions [28] in the genome (https://genome.sph.umich.edu/wiki/Regions_of_high_linkage_disequilibrium_(LD)) were removed. We used the RadialMR [29] package, which identify outlying genetic instruments via modified Q-statistics, to further exclude outlying pleiotropic SNPs. We also looked up each IV and its proxies (r^2^ > 0.8) in the PhenoScanner GWAS database (http://phenoscanner.medschl.cam.ac.uk) [30, 31] to assess any associations with potential confounders (smoking, drinking behavior, education, and socioeconomic status) or the outcome. SNPs associated with potential confounders or the outcome were removed. The remaining SNPs were used to perform MR analysis.

#### Testing IV strength and statistical power

We calculated the F-statistics to assess instrument strength [32]. IVs with F-statistics ≥ 10 indicates a relatively low risk of weak instrument bias in MR analysis [32]. Using the method proposed by Burgess [33], we also calculated the statistical power of our study. Briefly, the power was calculated by using the GWAS sample size, the proportion of cases in case–control GWASs and the variance explained by IVs for the exposure.

#### Bidirectional two-sample MR analyses

Two-sample MR analyses were performed to explore the bidirectional causal relationships between IDPs and T2D-related traits. The inverse variance weighted (IVW) method [34], which meta-analyzes the SNP specific Wald estimates using multiplicative random effects, was considered as the primary MR method. If the balanced pleiotropy assumption is violated, the causal estimates of IVW will be biased. Therefore, four other complementary MR methods were conducted to enhance the reliability of the results. MR-RAPS accounts for systematic and idiosyncratic pleiotropy and can provide a robust causal effect inference with many weak instruments [35]. MR-Egger method[34] is based on the INSIDE (instrument strength independent of the direct effects) assumption and estimates the causal effect through the slope coefficient of the Egger regression. The weighted median method assumes that at least 50% of the total weight of the instrument comes from valid variants [36] and estimates the causal effect from the median of the weighted empirical density function of individual IV effect estimates. The weighted mode method assumes that the most common causal effect is consistent with the true causal effect and provides a consistent effect estimate when the largest number of similar IV estimates come from valid instruments [37]. When only one IV was available, we used the Wald ratio[38] for MR analysis. The effect estimates are reported in β values for continuous outcomes and OR values for dichotomous outcomes. The analyses of all above MR methods were carried out using the TwoSampleMR package (v0.4.26) in R [39].

#### Sensitivity analysis

For the significant MR results after multiple testing correction, we carried out sensitivity analysis. First, leave-one-out analysis was carried out to check whether the causal association was driven by a single SNP. Second, the intercept term of the MR-Egger method was used to estimate the directional pleiotropic effect [40] (*P* < 0.05). Third, MR pleiotropy residual sum and outlier (MR-PRESSO) global test [41] was also used to detect horizontal pleiotropy (*P* < 0.05).

#### Data availability

The GWAS summary data of brain IDPs were obtained from the Oxford Brain Imaging Genetics web browser (https://open.win.ox.ac.uk/ukbiobank/big40/). The UK Biobank phenotypic data were obtained under the application number 46387. The GWAS summary data of T2D was downloaded from the DIAGRAM consortium web site (https://diagram-consortium.org/downloads.html). The summary data for the rest four traits (FG, 2hGlu, HbA1c, and FI) were obtained from the MAGIC web site (https://magicinvestigators.org/downloads/).

#### Patient and public involvement

This study did not involve patients or the public as it uses previously released GWAS summary data. Therefore, no patients or member of the public were involved in the design, or conduct, or reporting, or dissemination plans of our research.

## Results

### Study overview

The outline of this study is shown in Fig 1. Briefly, referring to our previous study [22], we obtained the GWAS summary data of 587 reliably measured IDPs derived from up to 33,224 European individuals of the UK Biobank (Figure 1). Summary data of five T2D-related traits were also collected (Supplementary Table S1). We manually checked the cohorts involved in these data and found that there was no sample overlap between the exposure-outcome pairs. Prior to MR analyses, genetic correlation analyses were performed and 142 pairs with *P* value < 0.05 (Supplementary Table S2) were detected. Then we conducted bidirectional two-sample MR analysis between these pairs and the significant threshold was set as *P* < 1.76 × 10^−4^ (0.05/142/2, 142 is the number of exposure-outcome pairs selected by genetic correlation analysis, 2 refers to forward and reverse MR tests).

### IV strength and statistical power

After selecting independent SNPs with linkage disequilibrium (LD) pruning, we removed the outlier IVs detected by RadialMR [29] (Supplementary Tables S3 and S4). IVs associated with potential confounders (Supplementary Tables S5 and S6) were also removed. For the final IVs, we calculated the F-statistics to assess instrument strength. As shown in Supplementary Tables S7 and S8, the F-statistics for the MR pairs with at least one IV were all over 30, indicating that the IVs are with suitable strength. The statistical power for each pair is also shown in Supplementary Tables S7 and S8. For example, we had 80% power at a significance level of 0.05 to detect a minimum OR > 1.06 or < 0.94 (Supplementary Table S7) when assessing the causal effect of IDPs on T2D, and a minimum β > 0.06 or < -0.06 when assessing the causal effect of T2D-related traits on IDPs (Supplementary Table S8). The power for significance level after multiple testing correction (1.76 × 10^−4^) is also listed in Supplementary Tables S7 and S8.

### The putative causal effects of IDPs on T2D-related traits

As shown in Figure 2 and Supplementary Table S9, we observed that 3 IDPs were causally associated with T2D or fasting insulin. The scatter plots are shown in Supplementary Figure S1.

**Figure 2.**
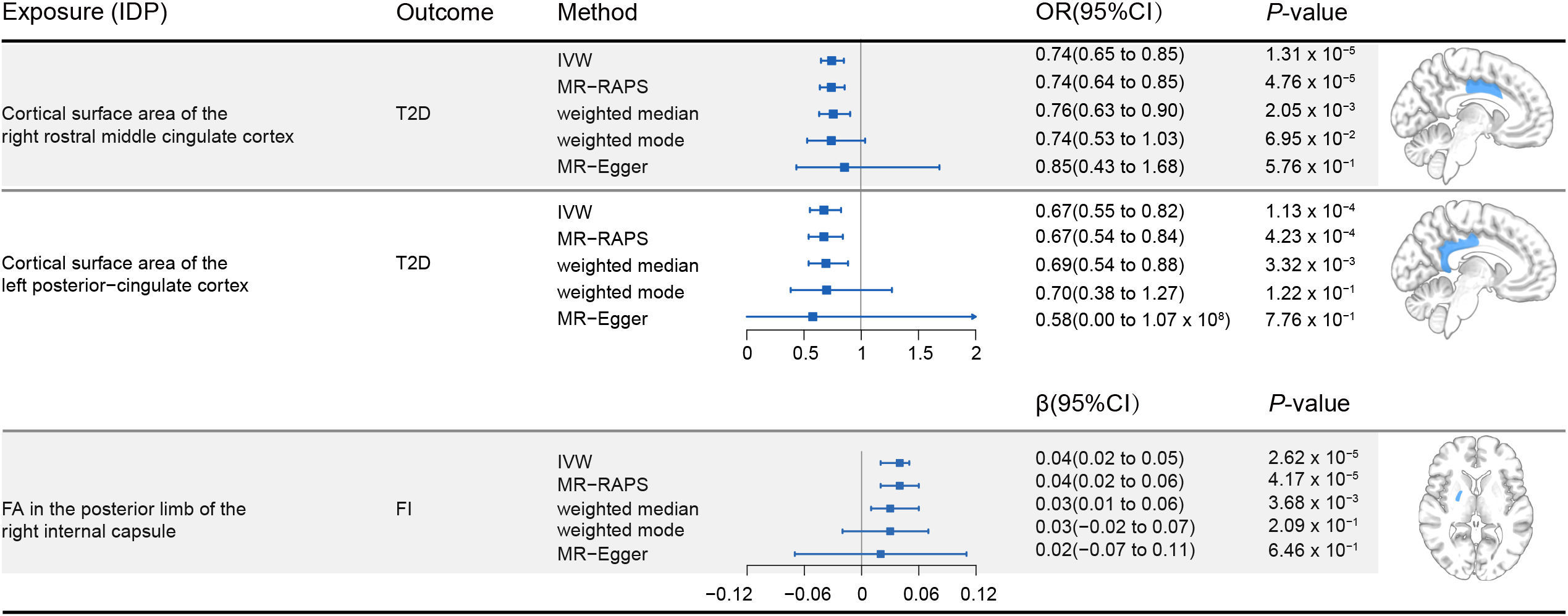
The putative causal effects of IDPs on T2D-related traits derived from Mendelian randomization analysis (inverse-variance weighted, MR-RAPS, weighted median, weighted mode and MR-Egger). Only results with significant inverse-variance weighted *P* value after multiple testing corrections are shown (*P* < 1.76 × 10^−4^). The diagram on the right shows the brain anatomical region of corresponding IDP. Abbreviations: T2D: Type 2 diabetes; IDP: Imaging-derived phenotype; FI: Fasting insulin.

Two IDPs were found to be associated with the risk of T2D. We observed that the area of the right rostral middle frontal cortex was negatively associated with the T2D risk (inverse-variance weighted (IVW) OR = 0.74, 95% CI 0.65 to 0.85, *P* = 1.31 × 10^−5^). The rostral middle frontal cortex, part of the dorsolateral prefrontal cortex, is a key brain region for emotion regulation and working memory [42]. We also found that the area of the left posterior cingulate cortex was negatively associated with the T2D risk (OR = 0.67, 95% CI 0.55 to 0.82, *P* = 1.13 × 10^−4^). The posterior cingulate cortex is a cortical hub of the human brain network [43], involving in several cognitive and affective functions, such as learning, memory, reward, and emotional stimulus processing [44].

For other T2D-related traits, we only found that fractional anisotropy (FA) in the posterior limb of the right internal capsule was positively associated with fasting insulin (β = 0.04, 95% CI 0.02 to 0.05, *P* = 2.62 × 10^−5^). The internal capsule is a white matter structure where many motor and sensory fibers travel to and from the cortex [45]. FA is a measurement of the degree of anisotropic diffusion occurring within a region (range from 0 to 1), it will be high in regions of high organization and approaching zero in free fluids. FA is widely used to reflect the structural integrity of the white matter [46].

### The putative causal effects of T2D-related traits on IDPs

As shown in Figure 3 and Supplementary Table S10, we observed that T2D-related traits might causally associated with 6 IDPs. The scatter plots are shown in Supplementary Figure S2.

**Figure 3.**
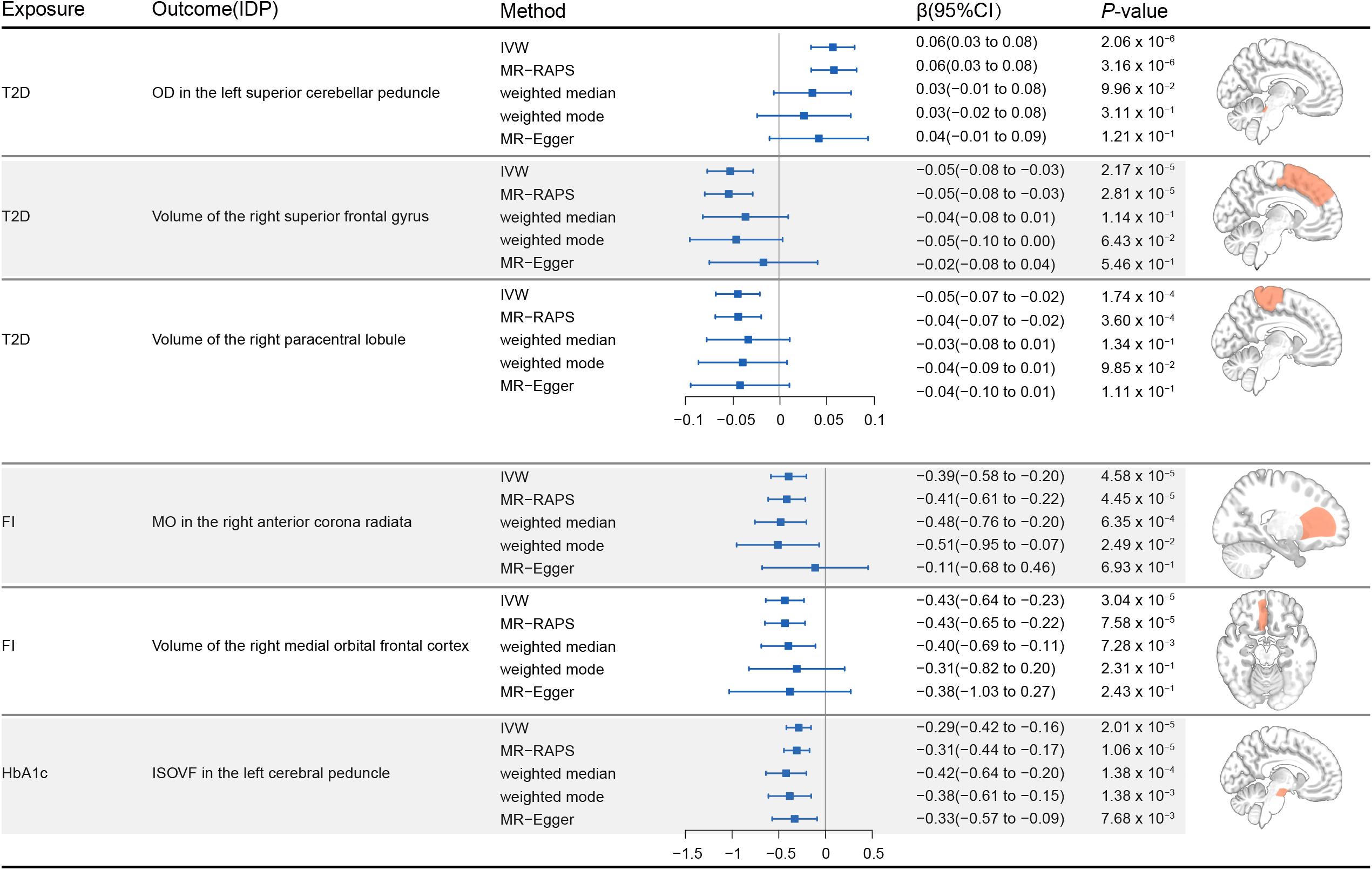
The putative causal effects of T2D-related traits on IDPs derived from Mendelian randomization analysis (inverse-variance weighted, MR-RAPS, weighted median, weighted mode and MR-Egger). Only results with significant inverse-variance weighted *P* value after multiple testing corrections are shown (*P* < 1.76 × 10^−4^). The diagram on the right shows the brain anatomical region of corresponding IDP. Abbreviations: T2D: Type 2 diabetes; IDP: Imaging-derived phenotype; FI: Fasting insulin. HbA1c: glycated hemoglobin.

T2D was found to be associated with 3 IDPs. We found that T2D was positively associated with orientation dispersion index (OD) in the left superior cerebellar peduncle (β = 0.06, 95% CI 0.03 to 0.08, *P* = 2.06 × 10^−6^). The superior cerebellar peduncle is a white matter structure involved in neural connectivity of the frontal–thalamic–cerebellar circuitry. OD is used to estimate the angular variation of neurites: ranging from 0 (all parallel) to 1 (isotropically randomly oriented) [47]. We found that T2D was negatively associated with the volume of the right superior frontal gyrus (β = -0.05, 95% CI -0.08 to -0.03, *P* = 2.17 × 10^−5^). The superior frontal gyrus is located at the superior part of the prefrontal cortex and has been reported to be involved in a variety of cognitive and motor control tasks [48]. We found that T2D was negatively associated with the volume of the right paracentral lobule (β = -0.05, 95% CI -0.07 to -0.02, *P* = 1.74 × 10^−4^). The paracentral lobule is located on the medial surface of the cerebral hemisphere, it controls motor and sensory innervations of the contralateral lower extremity.

For other T2D-related traits, fasting insulin and HbA1c were found to be associated with two IDPs and one IDP, respectively (Figure 3). We found that fasting insulin was negatively associated with the volume of the right medial orbital frontal cortex (β = -0.43, 95% CI -0.64 to -0.23, *P* = 3.04 × 10^−5^). The medial orbitofrontal cortex is subregion of the ventromedial prefrontal cortex and it has been implicated in goal-directed decision-making, reward representation, and emotional processing [49]. Fasting insulin was also found to be negatively associated with the diffusion tensor mode (MO) in the right anterior corona radiata (β = -0.39, 95% CI -0.58 to -0.20, *P* = 4.58 × 10^−5^). The anterior corona radiata is part of the limbic-thalamo-cortical circuitry and has been associated with emotion regulation and executive control of attention [50]. MO is mathematically orthogonal to FA, and it specifies the type of anisotropy, ranging from planar to linear [51]. We found that HbA1c was negatively associated with isotropic or free water volume fraction (ISOVF) in the left cerebral peduncle (β = -0.29, 95% CI -0.42 to -0.16, *P* = 2.01 × 10^−5^). The cerebral peduncle is the anterior part of the midbrain that connects the remainder of the brainstem to the thalami. The cerebral peduncles are the main highway for transporting signals from the cortex to other parts of the central nervous system, and are very important for body coordination. ISOVF is a measure of volume fraction of extracellular isotropic free water. An increased ISOVF might indicate an increase in extracellular isotropic fluid, such as in the case of neuroinflammation [47].

### Sensitivity analyses

We performed sensitivity analyses to validate the robustness of the causal associations we detected from the above bidirectional MR analyses. Leave-one-out analyses showed that no single SNP was driving the causal estimates (Supplementary Figures S3 and S4). In addition, both the MR-PRESSO global test and MR-Egger didn’t detect any significant evidence of horizontal pleiotropy (Supplementary Tables S11 and S12). The directions of the estimates from the four other complementary MR methods were the same to those from the IVW method. However, using the same significant threshold of *P* < 1.76 × 10^−4^ after multiple testing corrections, the numbers of associations supported by the four methods were less than the IVW method (Figure 2 and Figure 3). This might be caused by the fact that the power of the IVW method is greater than that of other four methods [37]. Therefore, the above sensitivity analyses confirmed the reliability of the causal associations we detected from bidirectional MR analyses.

## Discussion

Previous observational studies have reported the associations between IDPs and T2D, however, whether the relationships are causal is uncertain. Here we performed bidirectional MR analysis to investigate the causal associations between 587 IDPs and 5 T2D-related traits. We identified 3 IDPs with potential causal effects on T2D or fasting insulin. In addition, we identified potential causal effects of T2D-related traits on 6 IDPs.

When assessing the causal effects of IDPs on T2D-related traits, we found that both the areas of right rostral middle frontal cortex and left posterior cingulate cortex were negatively associated with the risk of T2D. Previous study has observed gray matter loss in medial frontal lobe and cingulate [16], our results further highlight the potential involvement of these two regions in the development of T2D. These two brain regions have been associated with emotion regulation [42, 43]. Previous longitudinal studies [52, 53] suggest that general emotional stress and anger are associated with an increased risk for T2D. The association between emotion regulation and T2D might be meditated by the corticotropin releasing factor - adrenocorticotropic hormone - cortisol pathway, which is activated in the stress response [54]. We also found that FA in the posterior limb of the right internal capsule was positively associated with fasting insulin. Evidence from observational studies between internal capsule and fasting insulin is still lacking. However, a previous study [55] showed that significantly decreased FA in internal capsule in T2D patients. Further studies are still needed to confirm the underlying mechanism.

When assessing the causal effects of T2D-related traits on IDPs, we found T2D was negatively associated with the volumes of the right superior frontal gyrus and the right paracentral lobule. Both the superior frontal gyrus and the anterior portion of the paracentral lobule belong to the frontal lobe. Fasting insulin was also found to be negatively associated with the volume of another subregion of the frontal lobe, the right medial orbital frontal cortex. Consistent with our results, several previous studies [56, 57] have observed reduced frontal lobe volume in diabetic patients when compared with controls. However, observational studies [58] which didn’t detect significant difference of frontal lobe volume between individuals with diabetes and controls also exist. Since observational studies can’t fully account for confounding factors, our results here may offer help to clarify the relationship between T2D and frontal lobe volume.

We also found that T2D was positively associated with OD in the left superior cerebellar peduncle. OD is a measure that reflects variations in neurite fiber orientations [47]. Although evidence from observational studies between T2D and OD of the cerebellar peduncle is lacking, FA of the cerebellar peduncle has been reported to be decreased in T2D patients. Fasting insulin was to be negatively associated with MO in the right anterior corona radiata. MO is mathematically orthogonal to FA, and it can also be used to evaluate white matter integrity [51].Observational studies between fasting insulin and MO in anterior corona radiata is lacking, but previous studies have observed reduced FA of anterior corona radiata in T2D patients. We found that HbA1c was negatively associated with ISOVF in the left cerebral peduncle. A previous study [59] showed decreased FA of cerebral peduncle in T2D patients with peripheral microvascular complications. Since ISOVF is a measure of volume fraction of extracellular isotropic free water, our results highlight the possibility abnormal HbA1c levels might affect the extracellular environment surrounding the neurites.

The limitations of this study should be addressed. First, since different ethnic populations have different LD structures and allele frequencies, IVs selected from data of complicated ethnic backgrounds may lead to biased causal estimates [60]. The GWAS summary data we used here were obtained from public database, therefore, we cannot assess the effects of population stratification on our results. However, we only used datasets derived from the European population in our analyses. Second, MR estimates the effect of a lifelong exposure on outcomes, which is different from the effect of an intervention during a specific duration [61]. Therefore, the estimated MR effect size is not equivalent to the effect size of randomized controlled trials for short-term interventions.

In conclusion, we performed bidirectional two-sample MR analyses to systematically investigate the causal associations between brain IDPs and T2D-related traits using large-scale GWAS summary data. Our results showed strong genetic evidence for causal links between neuroimaging phenotypes and T2D-related traits. The findings offer new insight into the pathophysiology and consequence of T2D.

## Data Availability

All data produced in the present work are contained in the manuscript

## Abbreviations

MR: Mendelian randomization
T2D: Type 2 diabetes
IDP: Imaging-derived phenotype
GWAS: genome-wide association studies
FG: fasting glucose
2hGlu: 2h-glucose post-challenge
HbA1c: glycated hemoglobin
FI: Fasting insulin
IV: Instrument variant
LD: Linkage disequilibrium
IVW: Inverse variance weighted
MR-PRESSO: MR pleiotropy residual sum and outlier

## Acknowledgement

We acknowledge the laboratories who released the GWAS summary data we used in public databases. We also thank the UK Biobank for developing and curating their data resources.

## Authors’ contributions

T.-L.Y. and S.-S.D. designed the study. S.-S.D. and Y.G. wrote and edited the manuscript. S.-S.D., K.Y., S.-H.T., and J.G. collected and analyzed the data. S.-S.D. and K.Y. drew the figures. T.-L.Y. had full access to all the data in the study and had final responsibility for the decision to submit for publication.

