## Supplementary for "Evaluation of the causal associations between brain imaging-derived phenotypes and type 2 diabetes: a bidirectional Mendelian randomization study"

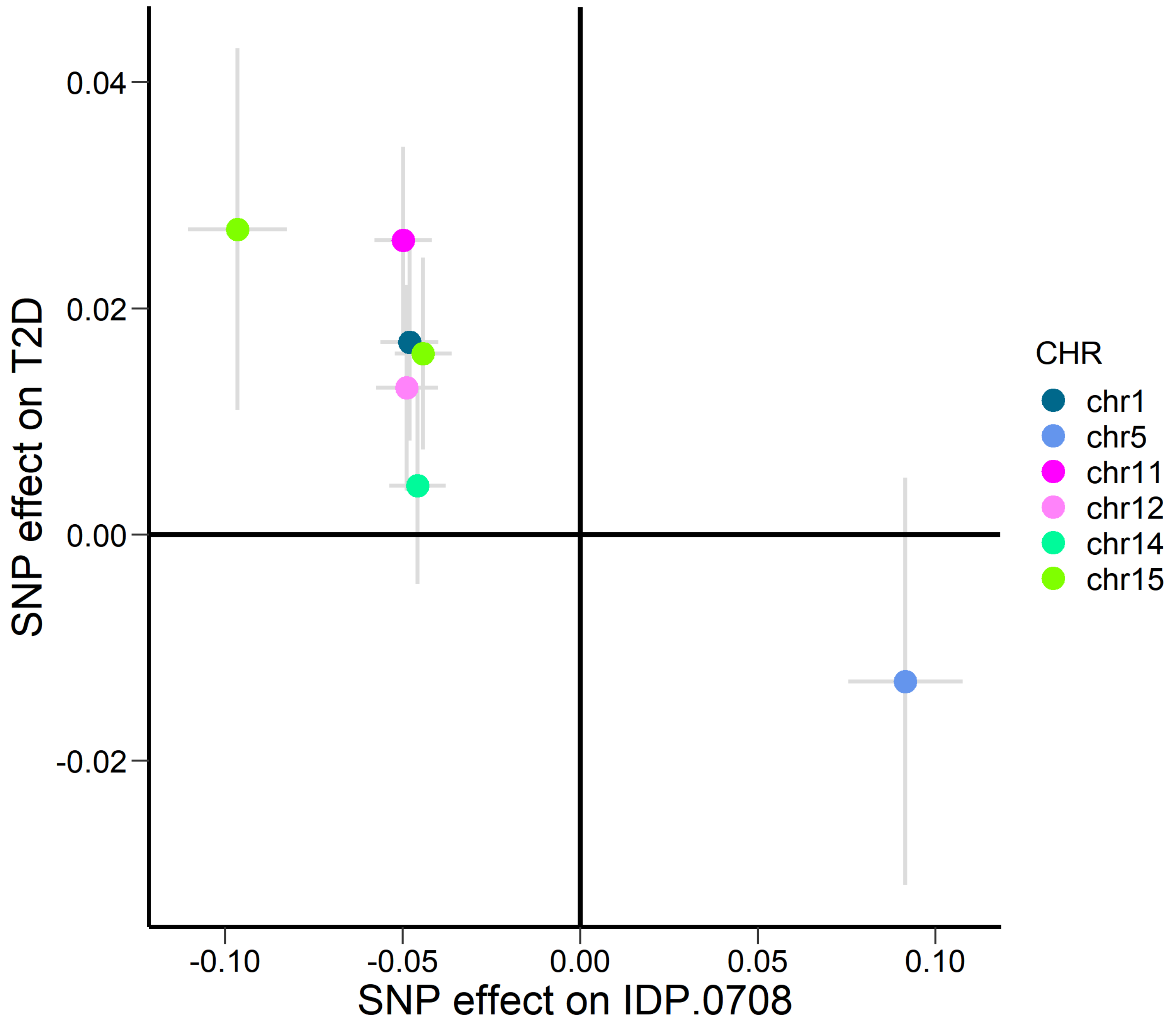


Scatter plot for cortical surface area of the right rostral middle cingulate on T2D

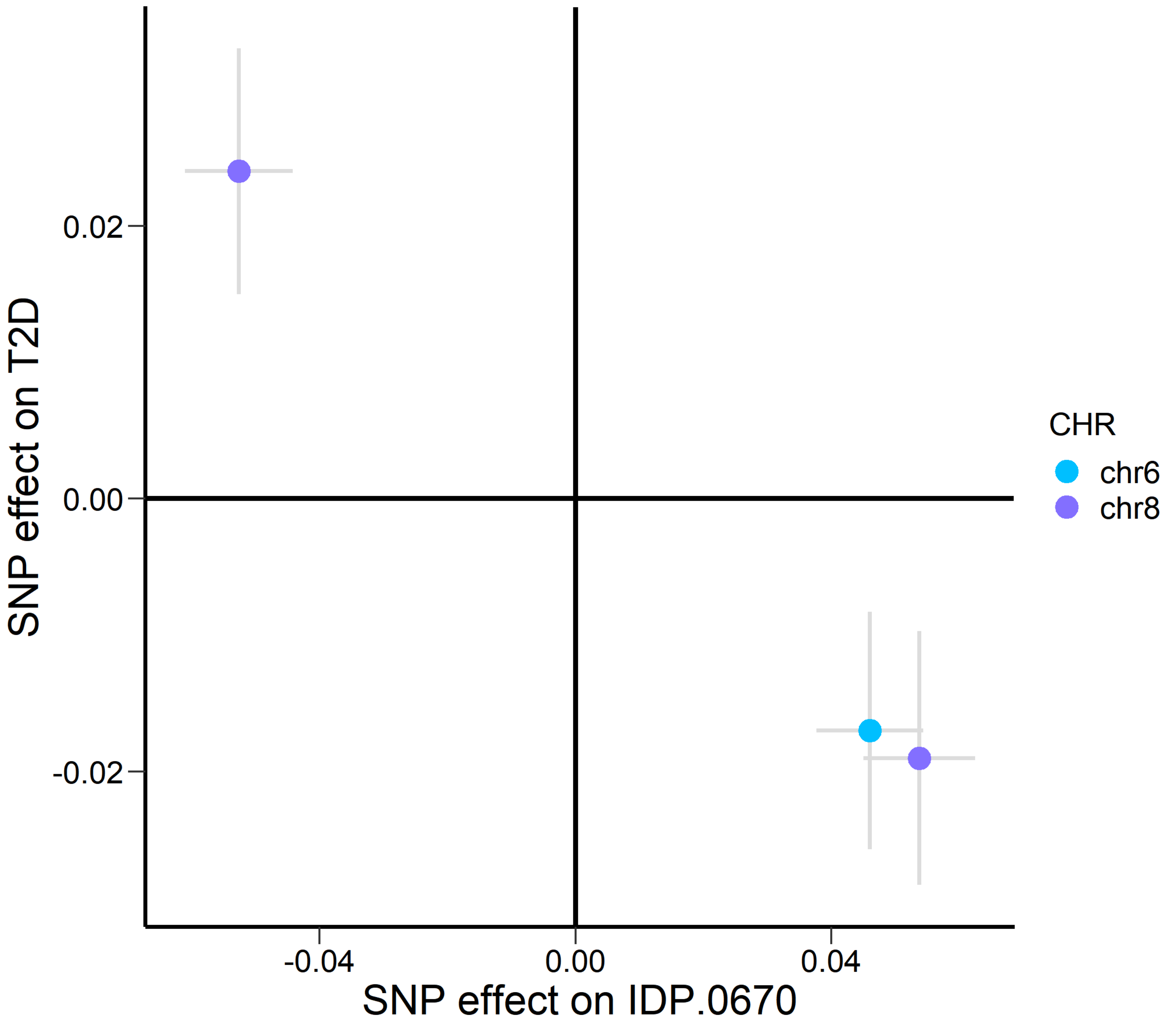


Scatter plot for cortical surface area of the left posterior-cingulate cortex on T2D

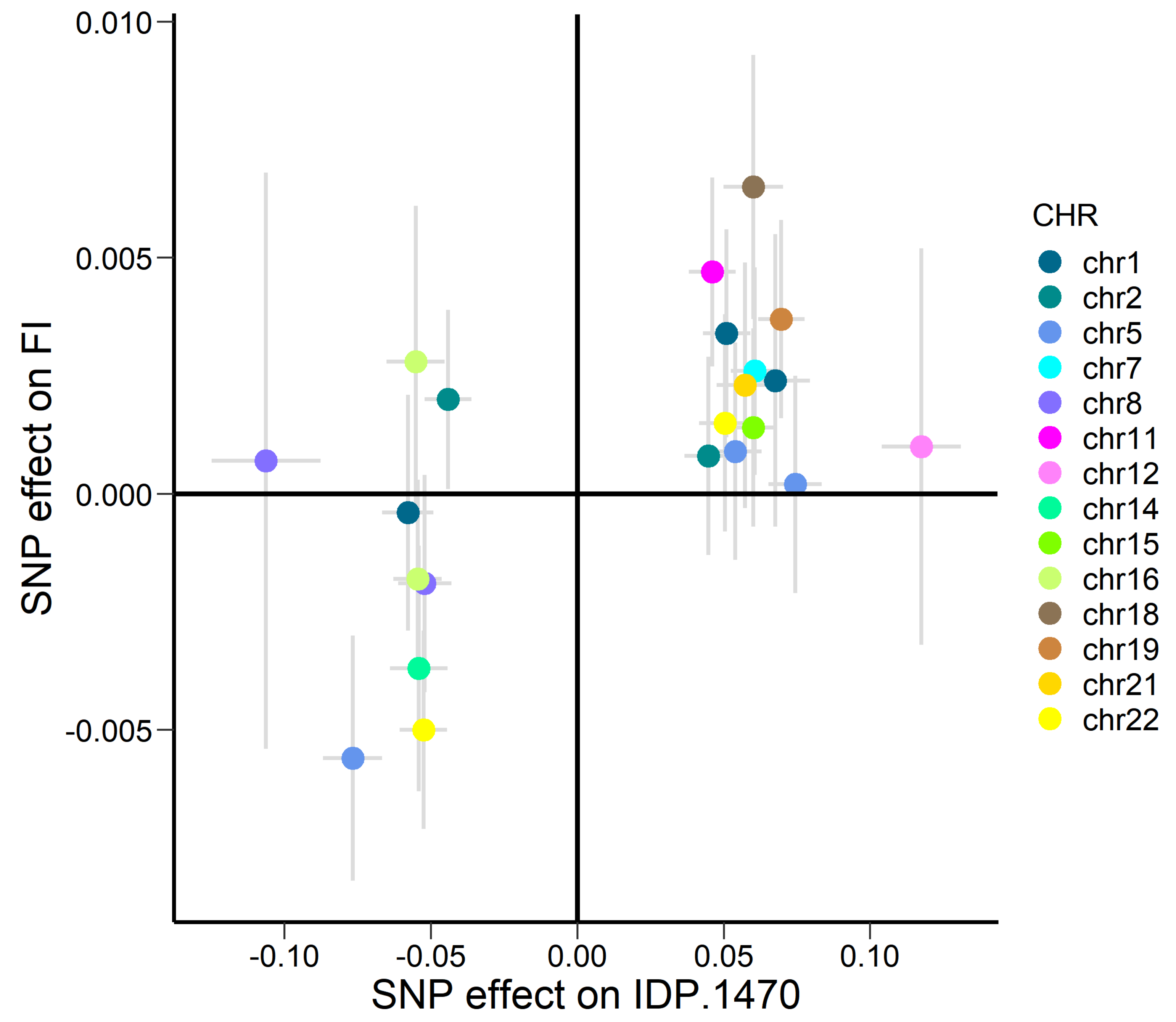


Scatter plot for FA in the posterior limb of the right internal capsule on fasting insulin

Figure S1. Scatter plot for the siginificant results when assessing the putative causal effects of IDPs on T2D-related traits.

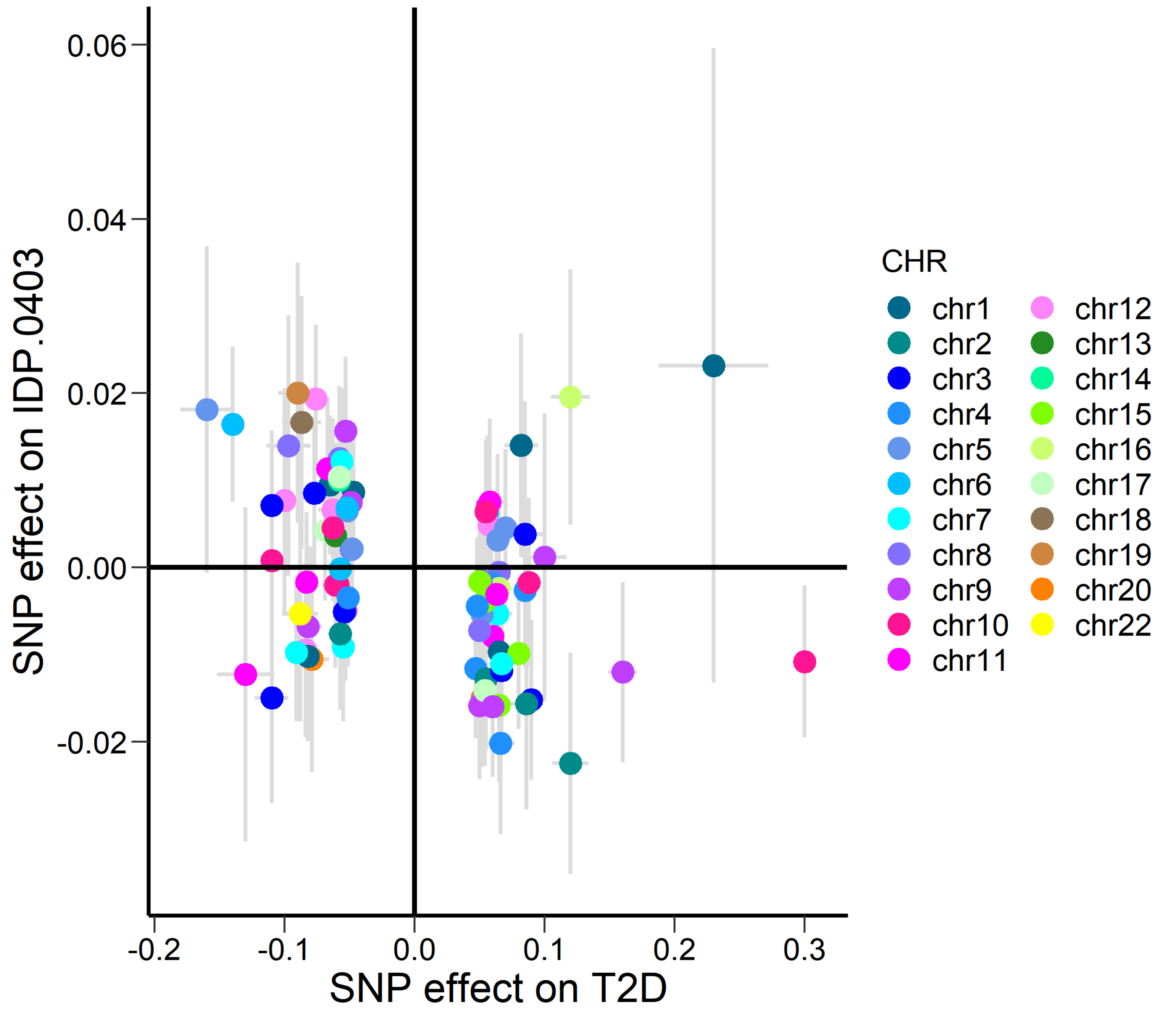


Scatter plot for T2D on volume of the right superior frontal gyrus

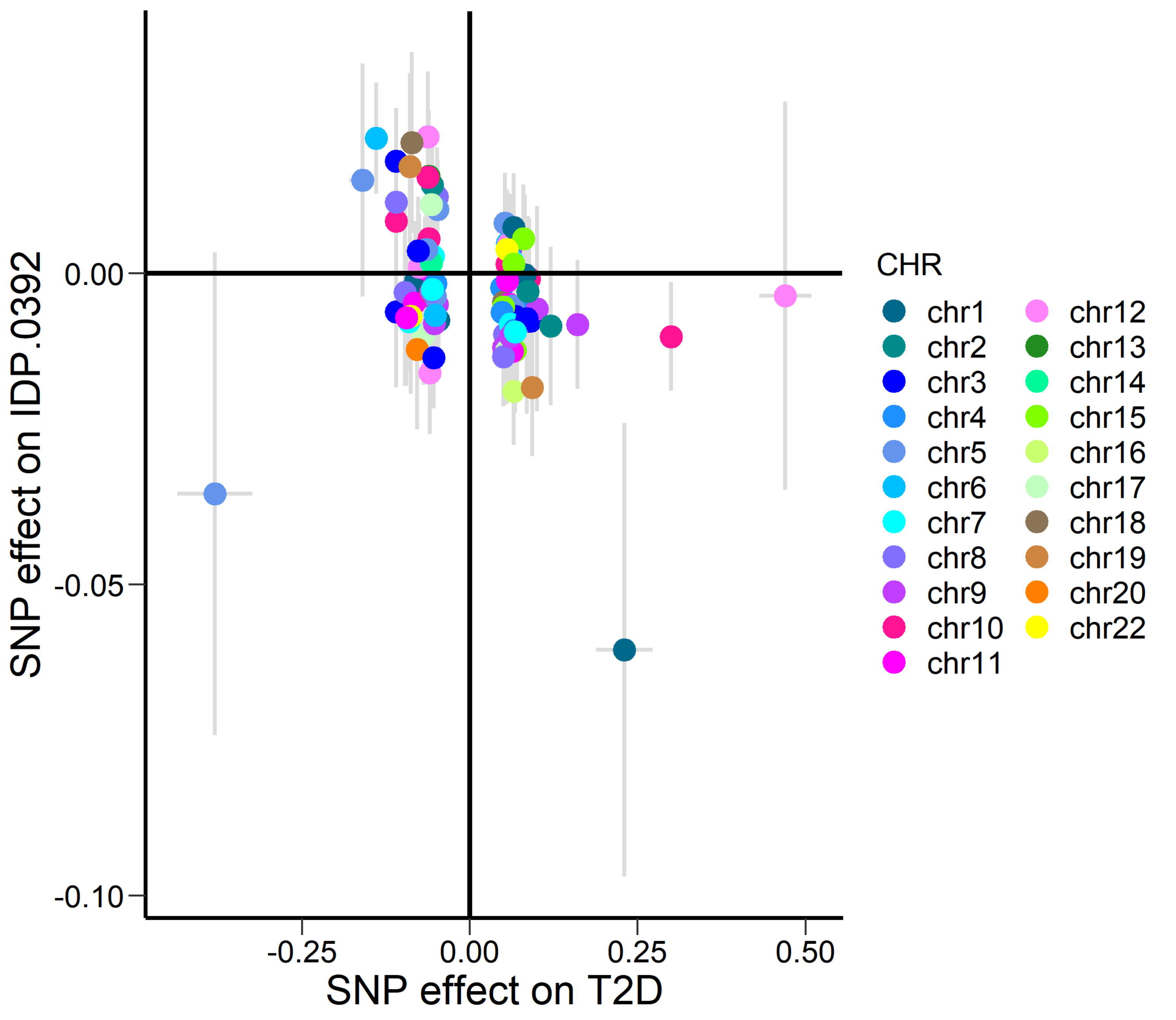


Scatter plot for T2D on volume of the right paracentral lobule

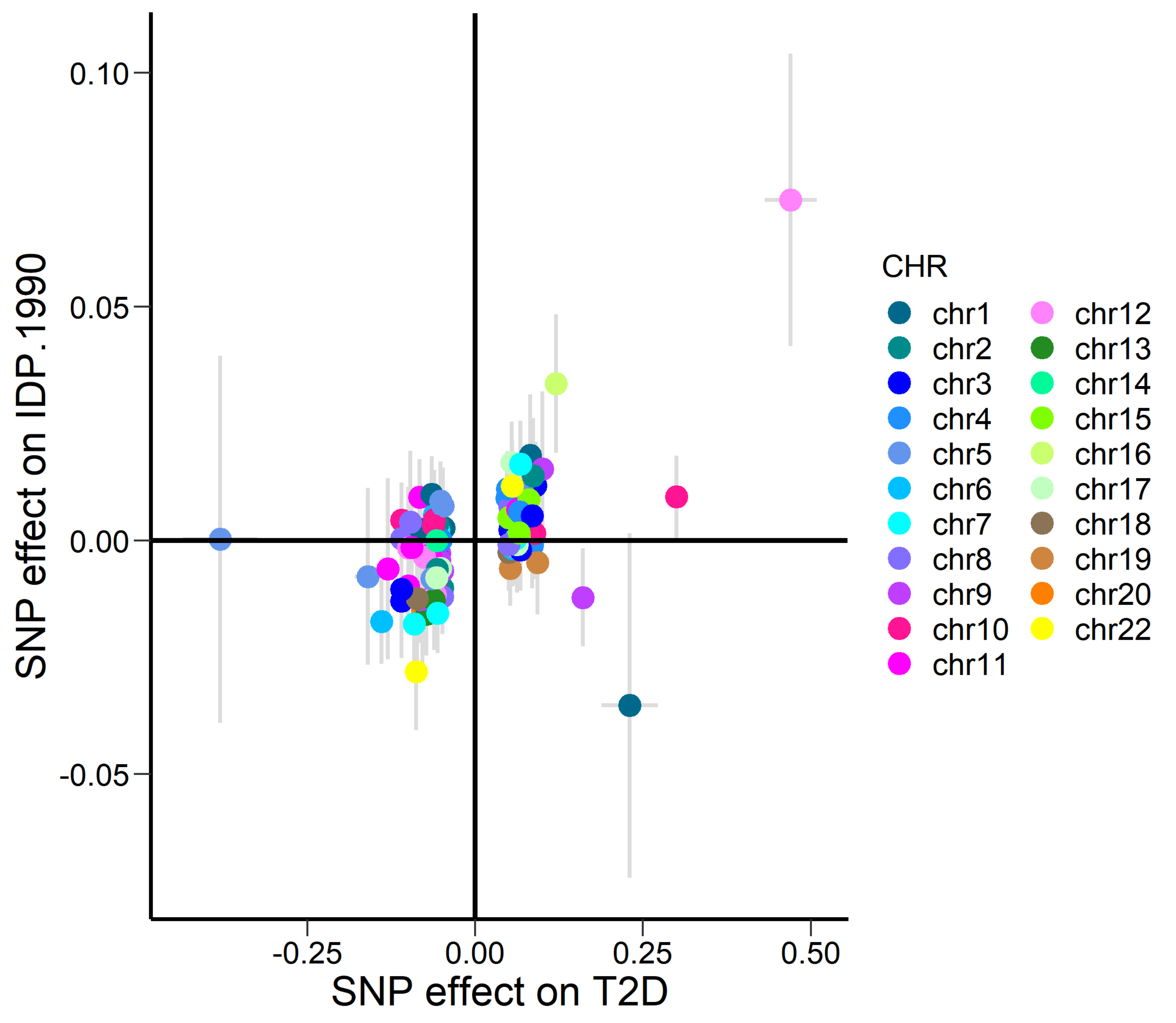


Scatter plot for T2D on OD in the left superior cerebellar peduncle

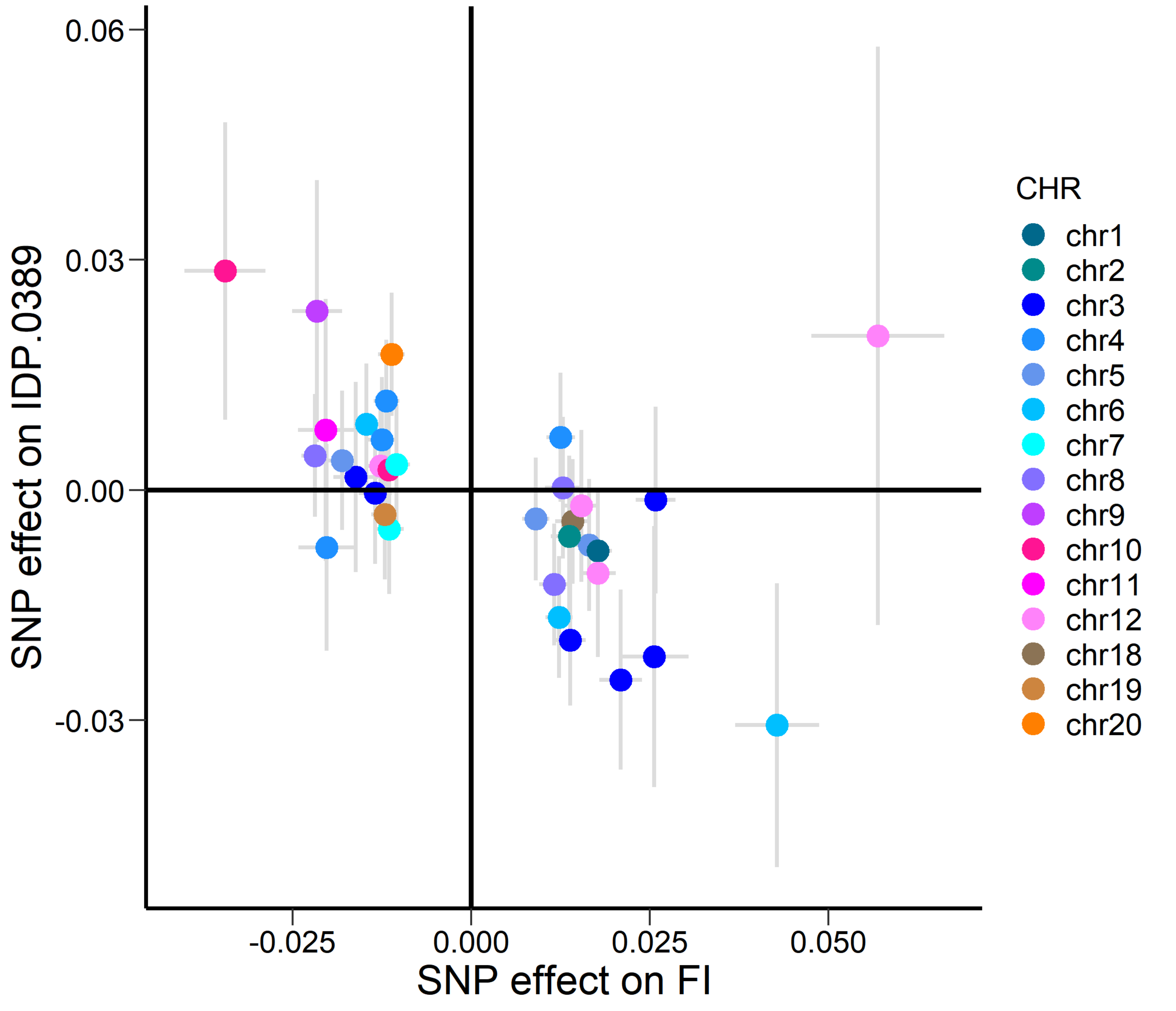


Scatter plot for fasting insulin on volume of the right medial orbital frontal cortex

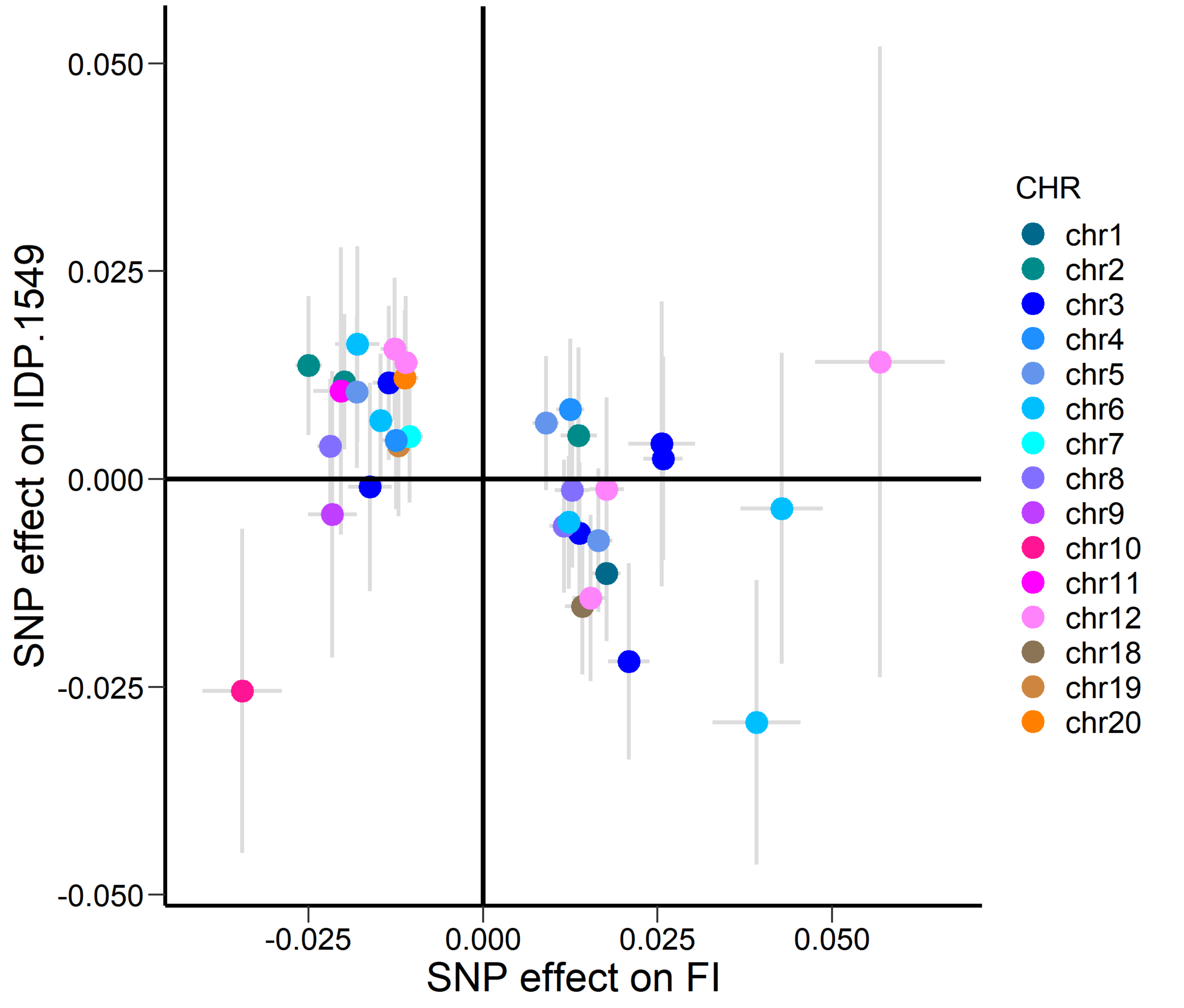


Scatter plot for fasting insulin on MO in the right anterior corona radiata

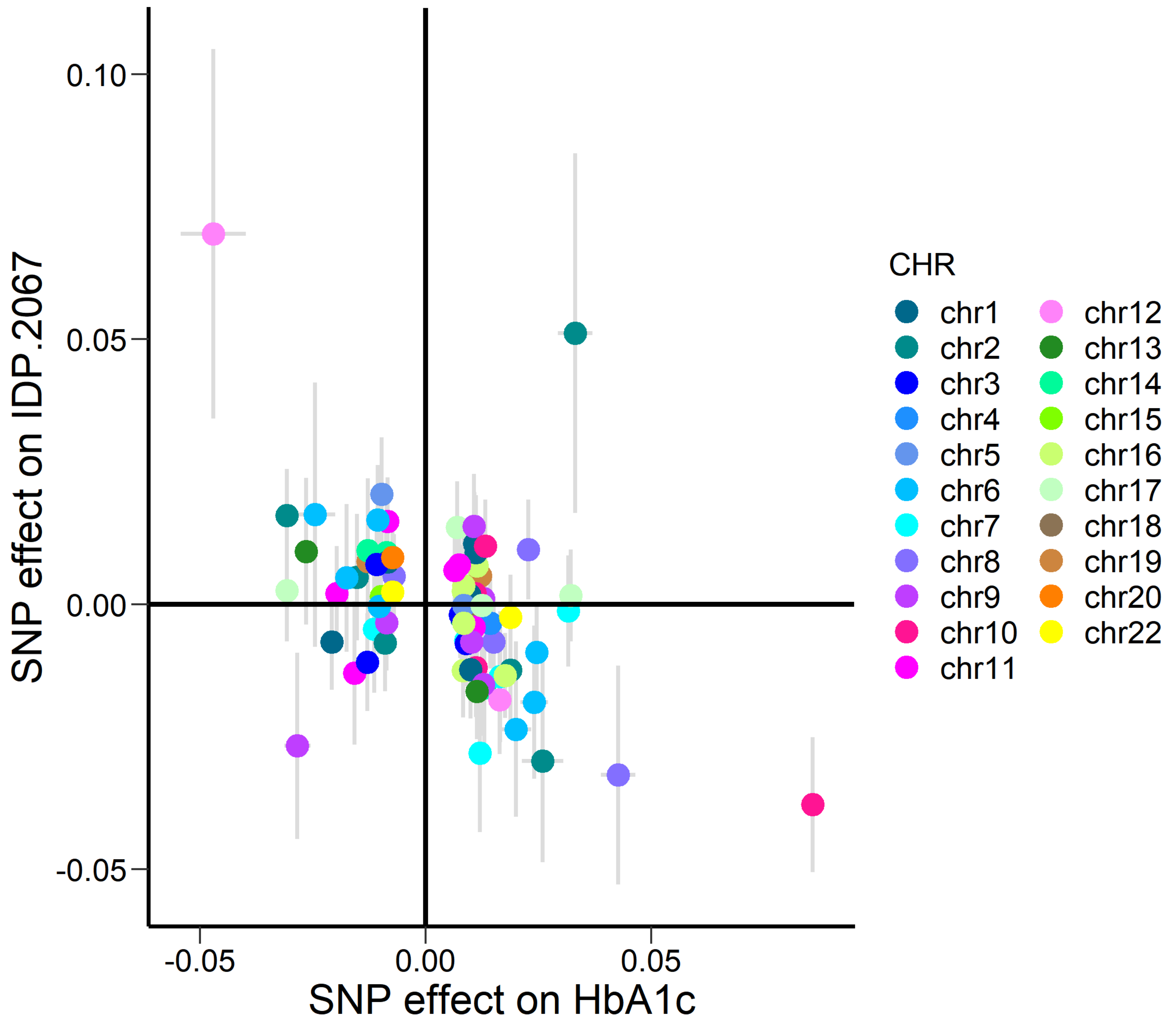


Scatter plot for HbA1c on ISOVF in the left cerebral peduncle

Figure S2. Scatter plot for the siginificant results when assessing the putative causal effects of T2D-related traits on IDPs.

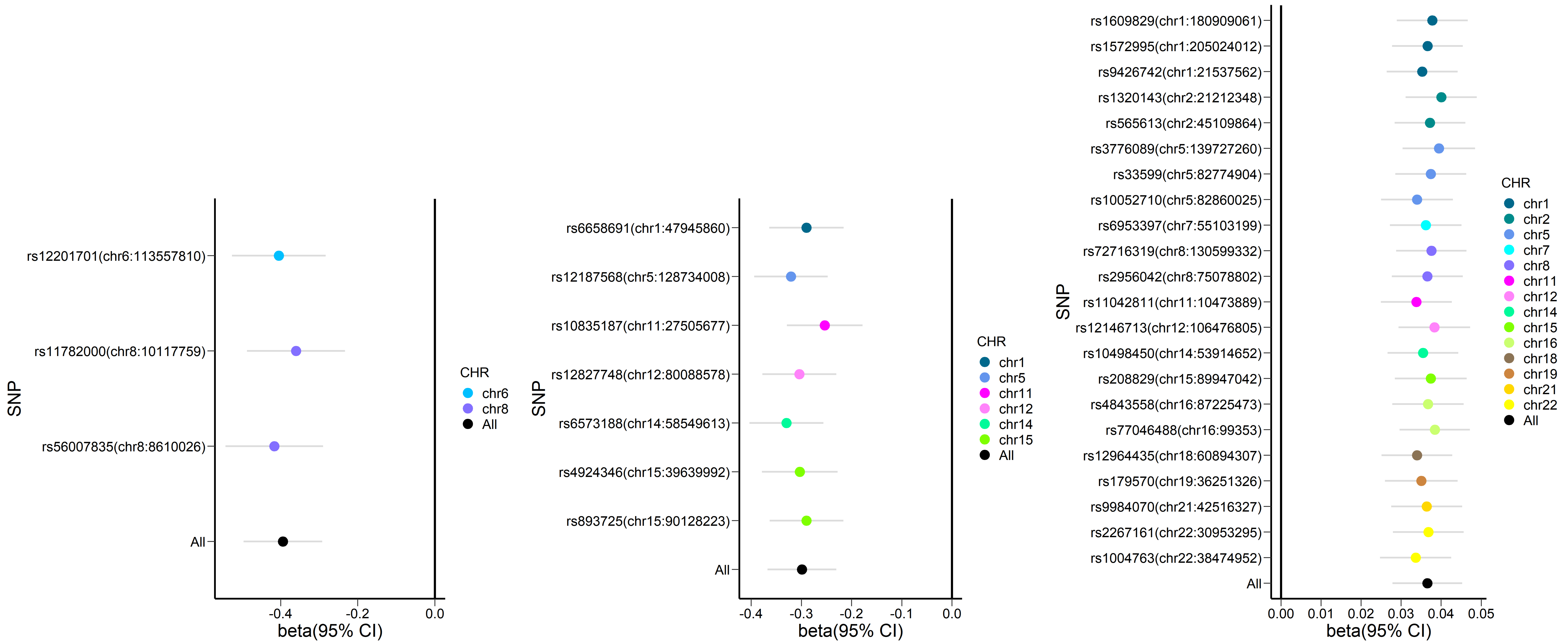


leave-one-out analysis for cortical surface area of the left posterior-cingulate cortex on T2D

leave-one-out analysis for cortical surface area of the right rostral middle cingulate on T2D

leave-one-out analysis for FA in the posterior limb of the right internal capsule on fasting insulin

Figure S3. Leave-one-out analysis plot for the siginificant results when assessing the putative causal effects of IDPs on T2D-related traits.

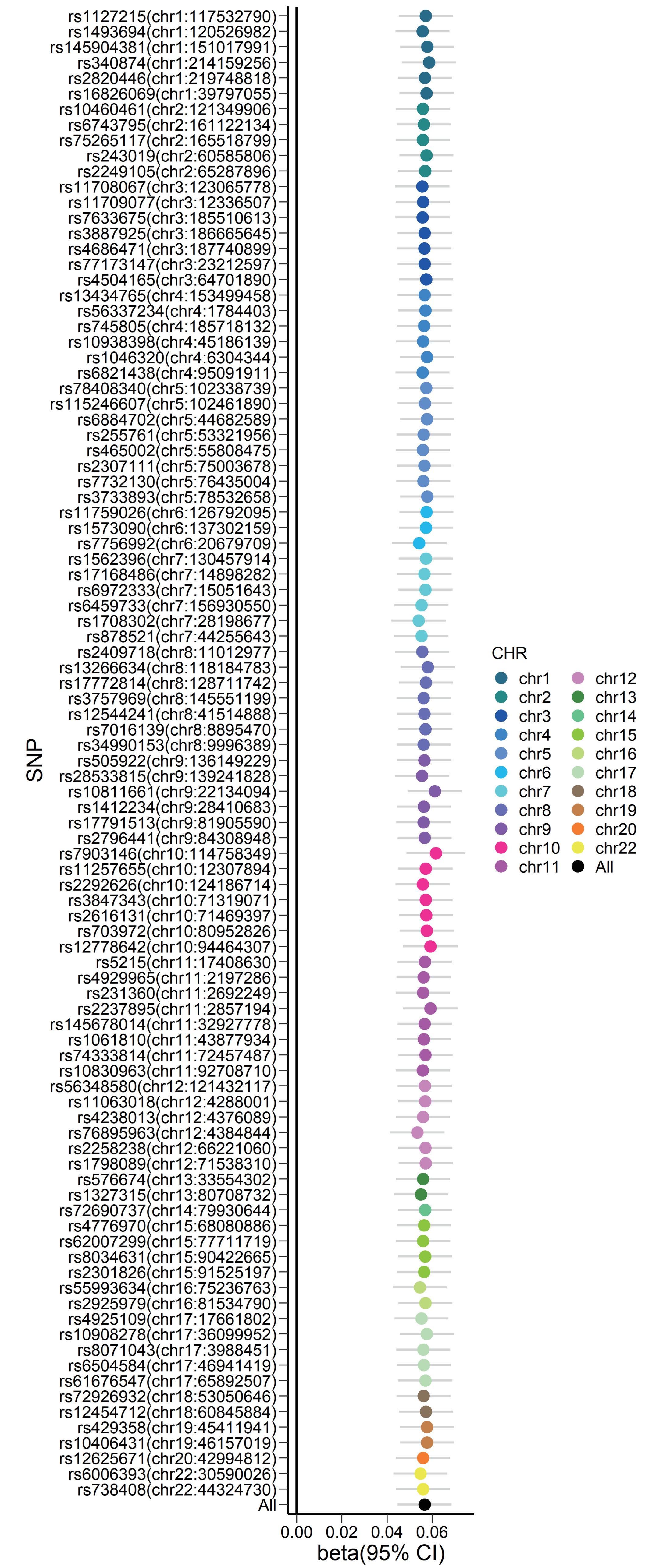


Leave-one-out analysis for T2D on OD in the left superior cerebellar peduncle

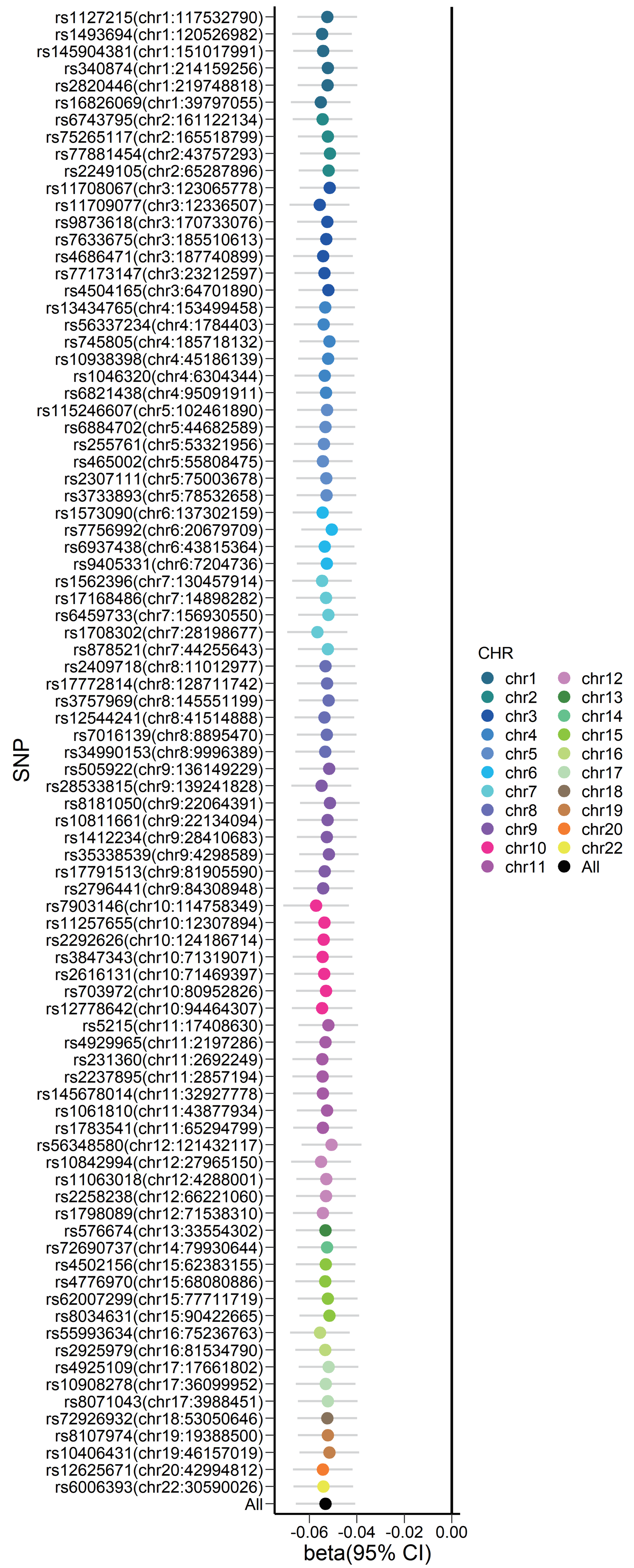


Leave-one-out analysis for T2D on volume of the right superior frontal gyrus

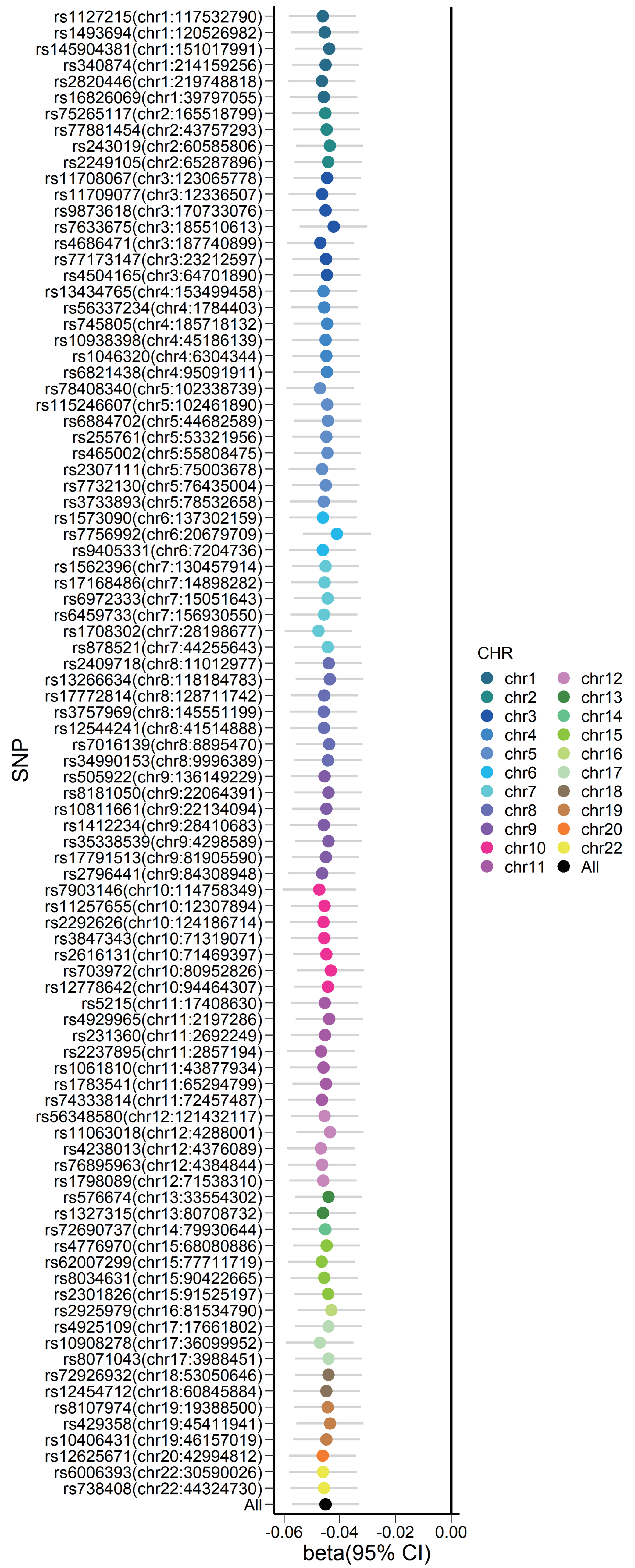


Leave-one-out analysis for T2D on volume of the right paracentral lobule

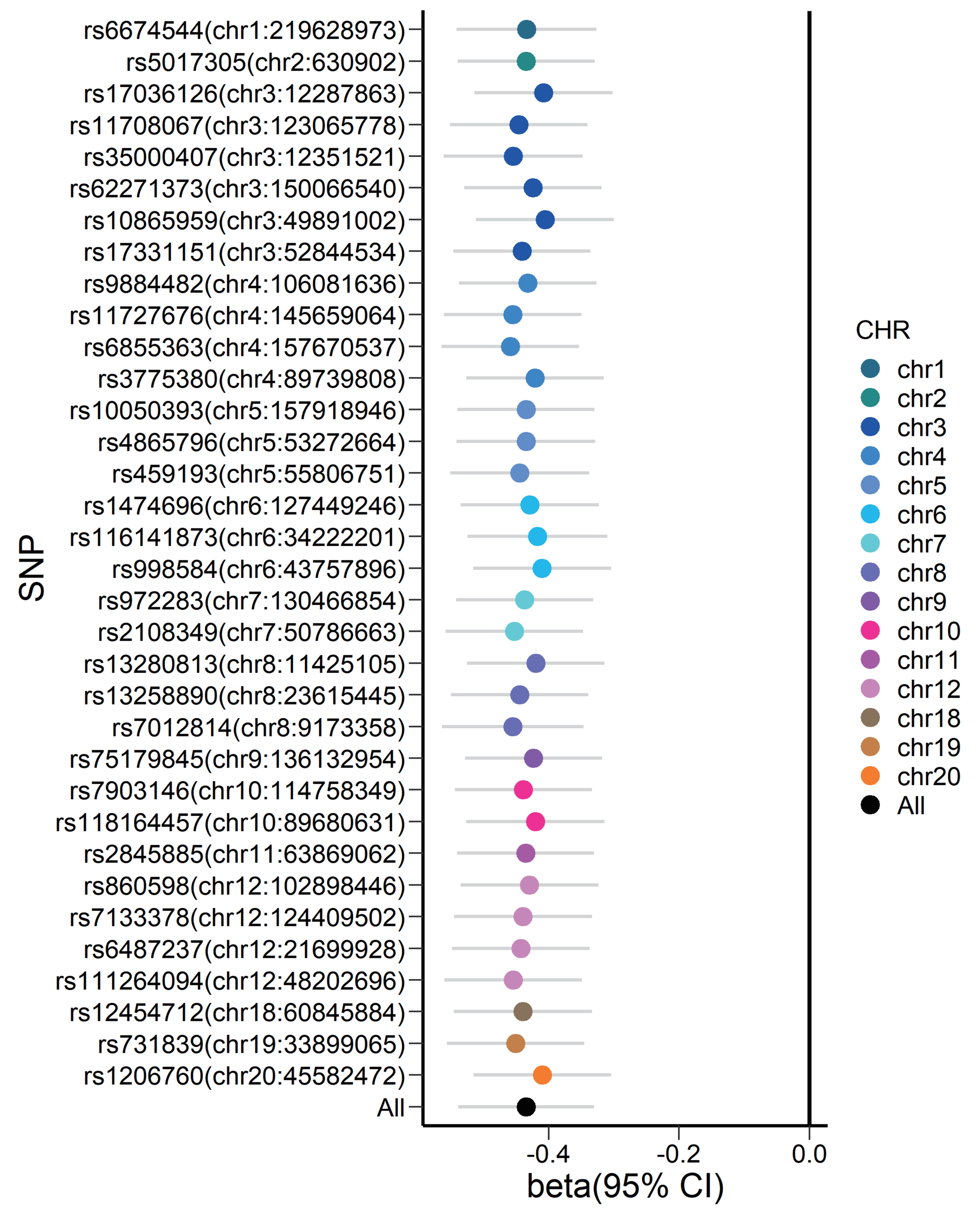


Leave-one-out analysis for fasting insulin on volume of the right medial orbital frontal cortex

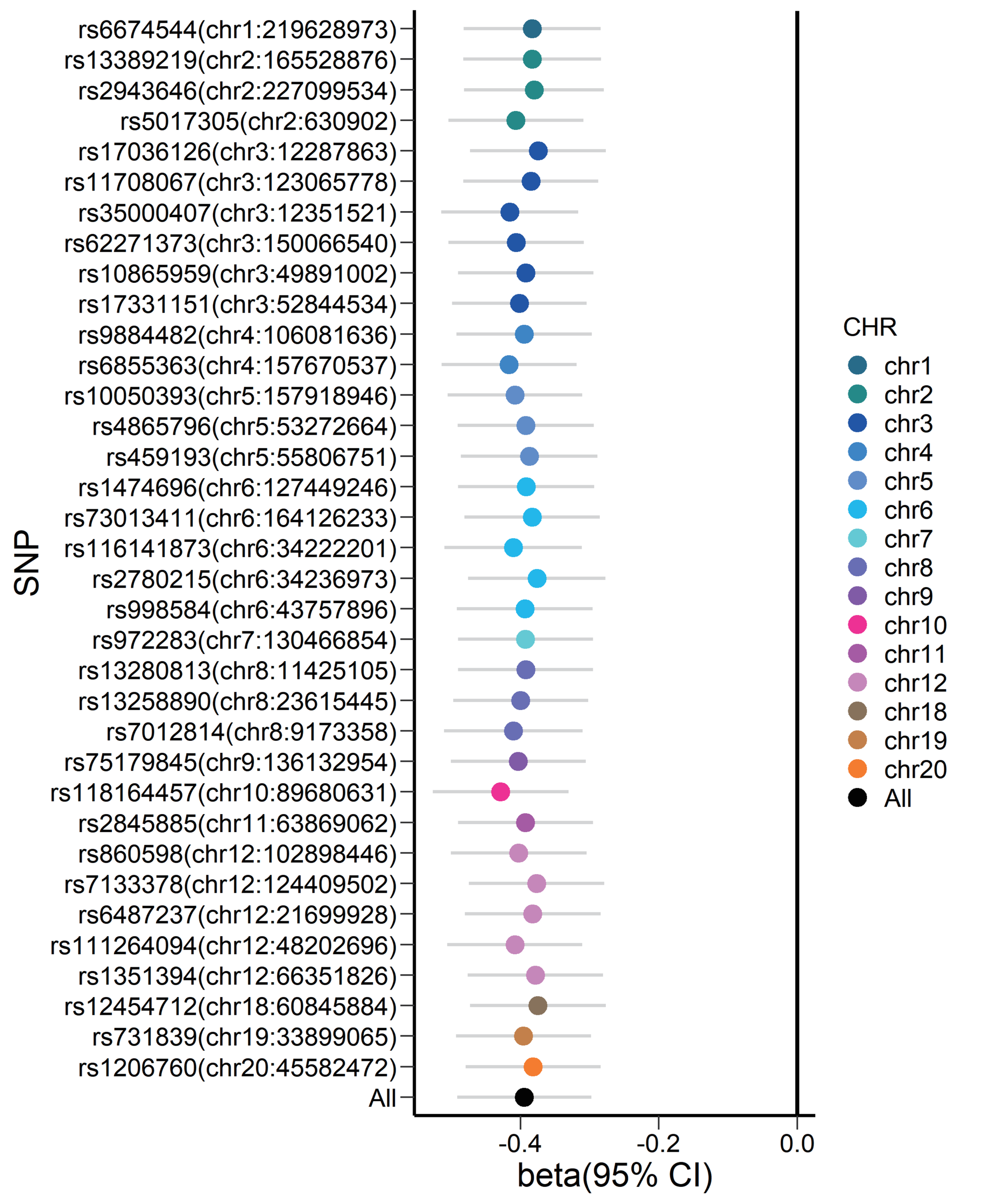


Leave-one-out analysis for fasting insulin on MO in the right anterior corona radiata

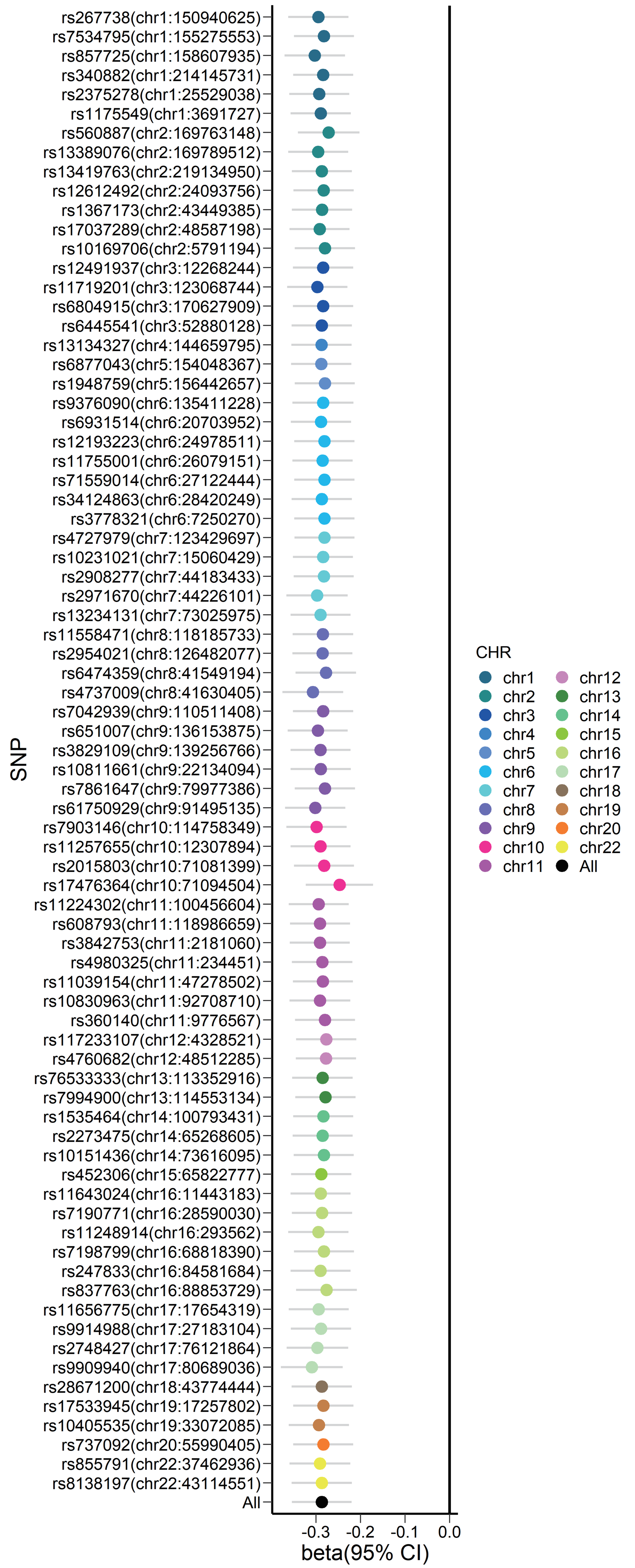


Leave-one-out analysis for HbA1c on ISOVF in the left cerebral peduncle

Figure S4. Leave-one-out analysis plot for the siginificant results when assessing the putative causal effects of T2D-related traits on IDPs.

**Table S1**. Information for the GWAS summary data used in this study.

| Trait | Abbreviation | Consortium | Max Sample size (cases) | PubMed ID | Download link |
|---|---|---|---|---|---|
| Brain imaging-derived phenotypes | IDP | UK Biobank | 33,224 | 33875891;36216997 | https://open.win.ox.ac.uk/ukbiobank/big40/ |
| Type 2 diabetes | T2D | DIAMANTE | 455313(55,005) | 30297969 | https://diagram-consortium.org/downloads.html |
| Fasting glucose | FG | MAGIC | 146,806 | 34059833 | https://magicinvestigators.org/downloads/ |
| 2h-glucose post-challenge | 2hGlu | MAGIC | 151,013 | 34059833 | https://magicinvestigators.org/downloads/ |
| Glycated haemoglobin | HbA1c | MAGIC | 200,622 | 34059833 | https://magicinvestigators.org/downloads/ |
| Fasting insulin | FI | MAGIC | 63,396 | 34059833 | https://magicinvestigators.org/downloads/ |

**Table S2**. Genetic correlation results between IDPs and T2D-related traits.

http://www.bigc.online/T2D_IDP/Table%20S2.xlsx

**Table S3**. Outliers detected by RadialMR when assessing the putative causal effects of IDPs on T2D-related traits.

http://www.bigc.online/T2D_IDP/Table%20S3-S6.xlsx

**Table S4**. Outliers detected by RadialMR when assessing the putative causal effects of T2D-related traits on IDPs.

http://www.bigc.online/T2D_IDP/Table%20S3-S6.xlsx

**Table S5**. IVs associated with potential confounders when assessing the putative causal effects of IDPs on T2D-related traits.

http://www.bigc.online/T2D_IDP/Table%20S3-S6.xlsx

**Table S6**. IVs associated with potential confounders when assessing the putative causal effects of T2D-related traits on IDPs.

http://www.bigc.online/T2D_IDP/Table%20S3-S6.xlsx

**Table S7**. The information of IVs for all exposure-outcome pairs when assessing the putative causal effects of IDPs on T2D-related traits.

http://www.bigc.online/T2D_IDP/Table%20S7-S8.xlsx

**Table S8**. The information of IVs for all exposure-outcome pairs when assessing the putative causal effects of T2D-related traits on IDPs.

http://www.bigc.online/T2D_IDP/Table%20S7-S8.xlsx

**Table S9**. The MR results when assessing the putative causal effects of IDPs on T2D-related traits.

http://www.bigc.online/T2D_IDP/Table%20S9-S10.xlsx

**Table S10**. The MR results when assessing the putative causal effects of T2D-related traits on IDPs.

http://www.bigc.online/T2D_IDP/Table%20S9-S10.xlsx

**Table S11**. Pleiotropy test for significant results when assessing the putative causal effects of IDPs on T2D-related traits.

| Exposure (IDP name) | IDP ID | Outcome | MR-PRESSO global test | | MR-Egger intercept | | |
|---|---|---|---|---|---|---|---|
| | | | **Observed RSS** | **P value** | **Effect size** | **SE** | **P value** |
| Cortical surface area of the right rostral middle cingulate cortex | IDP.0708 | T2D | 5.2832 | 0.7347 | -0.0078 | 0.0142 | 0.60 |
| FA in the posterior limb of the right internal capsule | IDP.1470 | FI | 20.0643 | 0.6482 | 0.0010 | 0.0024 | 0.68 |
| Cortical surface area of the left posterior-cingulate cortex | IDP.0670 | T2D | - | - | 0.0080 | 0.0761 | 0.93 |

Note: T2D: Type 2 diabetes; FI: Fasting insulin.

**Table S12**. Pleiotropy test for significant results when assessing the putative causal effects of T2D-related traits on IDPs.

| Exposure | Outcome (IDP name) | IDP ID | MR-PRESSO global test | | MR-Egger intercept | | |
|---|---|---|---|---|---|---|---|
| | | | **Observed RSS** | **P value** | **Effect size** | **SE** | **P value** |
| T2D | OD in the left superior cerebellar peduncle | IDP.1990 | 66.95 | 0.99 | 0.0013 | 0.0021 | 0.53 |
| HbA1c | ISOVF in the left cerebral peduncle | IDP.2067 | 77.55 | 0.50 | 0.0009 | 0.0020 | 0.67 |
| T2D | Volume of the right superior frontal gyrus | IDP.0403 | 78.57 | 0.76 | -0.0031 | 0.0023 | 0.18 |
| FI | Volume of the right medial orbital frontal cortex | IDP.0389 | 24.07 | 0.92 | -0.0009 | 0.0050 | 0.86 |
| FI | MO in the right anterior corona radiata | IDP.1549 | 25.26 | 0.91 | -0.0051 | 0.0047 | 0.29 |
| T2D | Volume of the right paracentral lobule | IDP.0392 | 79.78 | 0.84 | -0.0002 | 0.0021 | 0.92 |

Note: T2D: Type 2 diabetes; FI: Fasting insulin; HbA1c: glycated hemoglobin.
